# Pancreatic cancer risk prediction using deep sequential modeling of longitudinal diagnostic and medication records

**DOI:** 10.1101/2025.03.03.25323240

**Authors:** Chunlei Zheng, Asif Khan, Daniel Ritter, Debora S. Marks, Nhan V. Do, Nathanael R. Fillmore, Chris Sander

**Author notes:** Joint first authors (Chunlei Zheng, Asif Khan, Daniel Ritter). Joint senior authors (Nathanael R. Fillmore, Chris Sander).

## Abstract

**Background:** Pancreatic ductal adenocarcinoma (PDAC) is a rare, aggressive cancer often diagnosed late with low survival rates, due to the lack of population-wide screening programs and the high cost of currently available early detection methods.

**Methods:** To facilitate earlier treatment, we developed an AI-based tool that predicts the risk of pancreatic cancer diagnosis within 6, 12 and 36 months of assessment, using time sequences of diagnostic and medication events from real-world electronic health records (EHRs). Trained on a large US Veterans Affairs dataset with 19,000 PDAC cases and millions of controls, the tool employs a Transformer-based model that can capture and benefit from information synergy between diagnoses and medications.

**Findings:** Risk prediction is improved when incorporating medication data alongside diagnostic codes. For N patients predicted to be at highest risk out of 1 million, risk of cancer within 3 years is substantially higher than using a reference estimate based on age and gender alone (standard incidence ratio SIR=115 to 70 for N=1000 to 5000). Detection of the most predictive features generates clinical hypotheses such as the role of chronic inflammatory conditions in predisposing to PDAC or use of specific medication that highlight the health state of a patient and cancer risk. We quantify prediction bias between different socioeconomic subpopulations.

**Interpretation:** The risk prediction tool is intended to be the first step in a three-step clinical program: identification of high-risk individuals using AI tools, followed by a stratified surveillance program for early detection and intervention, aiming to benefit patients and lower health-care costs.

**Funding:** US CDMRP Pancreatic Cancer Risk Using Artificial Intelligence.

## Introduction

PDAC is one of the most challenging cancers to diagnose with a poor prognosis, often detected at advanced stages when curative treatment options are limited. Its increasing incidence makes it one of the leading causes of cancer-related deaths worldwide ^(1),(2)^. Approximately 80% of pancreatic cancer patients are diagnosed with locally advanced or distant metastatic disease and have poor survival rates (only ∼9% survive for 5 years) (3). In contrast, for the ∼20% of patients with early stage disease (stage IA) survival rates are much higher (∼80% of these survive for 5 years, according to the US Veterans Affairs (US-VA) cancer registry (4)). These patients can be effectively treated by a combination of surgery, chemotherapy and radiotherapy (5). Therefore, identifying risk factors and improved detection at early stages could significantly improve outcomes and reduce mortality from this aggressive malignancy.

### Early risk models on selected cohorts

Early detection programs can improve patient outcomes, but universal implementation is currently impractical due to the risk of false positives and high cost of screening for some cancer types, such as PDAC. An affordable selective screening approach targets high-risk individuals with an acceptable low rate of false positives. Previous works on selected (but relatively small) cohorts have developed risk models using factors such as family history, lifestyle, environmental, and genetic variants (6–11), and identified individuals at increased risk of PDAC when such data was available (12).

### Using real-world EHR data in predictive AI models

Electronic health records (EHRs) offer a rich and generally available source of patient data, including diagnoses, medications, and laboratory results, that can support population-wide risk prediction. Prior works have demonstrated the application of machine learning (ML) in EHR systems, such as BEHRT, a Transformer model specifically designed for general prediction of future diagnoses of patients (13). Similar ML approaches have been explored in other recent works (14–17). Training specific ML prediction models on routinely collected large-scale real-world EHR data could enable targeted screening and preventive interventions for high risk PDAC patients. Recent works have used EHR data to develop several PDAC risk prediction models (3);(18–22), which can be used to identify high risk cohorts.

### Medication data for improved risk prediction

Most cancer risk prediction models based on EHR data predominantly rely on diagnosis codes alone and only a few use other forms of data types, such as medication records (3,19,21). Diagnosis codes, especially in the US healthcare system, are influenced by billing incentives and reimbursement practices, which can lead to “billing pollution”, where differential or secondary diagnosis codes are upcoded to justify costly procedures or maximize reimbursement (23,24). This data noise can result in a somewhat inaccurate representation of a patient’s state of health. In contrast, medication data plausibly provide more objective information as prescriptions are based on well-trained physicians’ decisions and are more likely to represent objective clinical necessity rather than financial motivation. In addition, use of medications can provide direct information on disease risk due to known preventive effects or negative side effects.

### Drug effects on cancer risk

The usage of certain medications has been linked to altered cancer risk,with either positive or negative effects. E.g., consider omeprazole, a proton pump inhibitor commonly used to treat gastroesophageal reflux disease. Recent epidemiological studies have raised concerns based on potential association between long-term use of omeprazole and an increased risk of developing PDAC (25–27). This association is attributed to hypergastrinemia, a condition characterized by elevated levels of the hormone gastrin, which has been shown to stimulate the growth of pancreatic cancer cells via interaction with the cholecystokinin B receptor (CCKBR). An opposite example is metformin, commonly used to treat type 2 diabetes, which has been associated with a decreased risk of PDAC (28). Metformin works by lowering blood glucose levels (29,30) and also reduces levels of inflammatory markers (31,32), both of which are factors that can contribute to cancer development. While such associations with altered risk may be influenced by various confounding factors, medications may provide non-redundant information about cancer risk.

### Non-sequential versus sequential risk models

Previous risk prediction efforts have primarily relied on a pre-identified set of diagnosis codes as input features, overlooking valuable longitudinal information within EHR and the sequential nature of clinical events. Recently, a PDAC RISk Model (‘PRism’) (3) trained on a large-scale TriNetX (33) database, incorporated composite features, including diagnosis records, medication history, and laboratory results to encode each patient’s history as a fixed 87-dimensional vector, which was then used to train multilayer perceptron (MLP) and logistic regression (LR) models. The choice of input features in PRism does not model the sequential ordering and timing of events explicitly. Unlike PRism, our AI tool uses deep sequential learning, explicitly capturing the sequential order and timing of clinical events for more nuanced PDAC risk prediction.

### Combination model trained on medication and diagnostic data

Previously, CancerRiskNet (34) introduced a Transformer-based sequential model for PDAC risk prediction, using only diagnosis codes. Here, we extend this architecture to longitudinal sequences of both diagnosis and medication codes, allowing the model to learn a shared encoded space that captures both clinical diagnostic histories and prescribed drugs over time. Our model utilizes self-attention to capture interactions among disease and medication events, while accounting for temporal spacing using positional encoding based on relative event timings. Developed and tested on a large-scale US-VA dataset, this model is a novel approach to integrate diagnostic and medication data for PDAC risk prediction. Our work facilitates identification of high risk cohorts who could benefit from a three step clinical program – prediction, detection, and intervention (Figure 1B), to improve patient outcomes.

**Figure 1:**
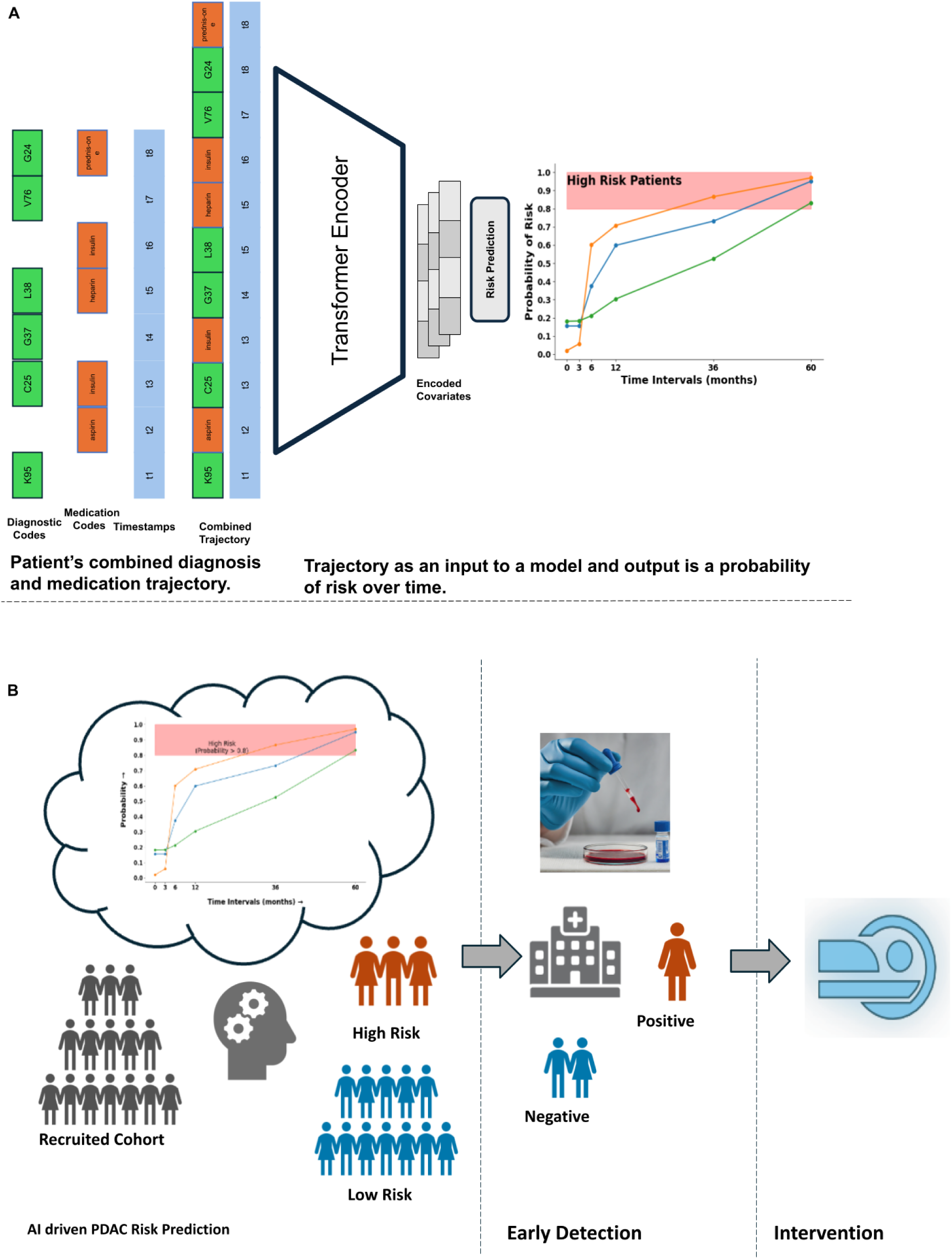
AI Tool for Risk Prediction. **A:** Example of input trajectory based on the combination of diagnostic (ICD) and medication (RxNorm) codes. A trajectory represents the patient’s medical history, encompassing diagnoses and prescribed medications along with respective timestamps. The combined trajectory is given as input to the AI prediction tool, which uses a Transformer encoder to extract latent risk factors. These factors are then fed to a risk prediction model that calculates the probability of cancer risk over different time intervals. **B:** Surveillance program that uses the AI tool to identify high-risk patients. Identified high risk cohorts can benefit from early detection technologies and possible healthcare interventions.

## Results

### Performance of combination model trained with data exclusion

The ICD codes appearing shortly before the diagnosis of cancer can represent quasi-symptoms that would be easily interpreted by a clinician. To determine whether the model is primarily based on these quasi-symptoms or learns non-trivial covariates predictive of cancer, we trained models with and without a three-month data exclusion window. The data exclusion involves removing all ICD codes and medications assigned within three months prior to the diagnosis of pancreatic cancer. With this data exclusion, compared to models from either ICD or medication alone, the combination model performed best (for each of the ‘cancer within k months’ intervals). For instance, for the combined model the AUROC is 0.857 (95% CI: 0.852-0.860) for the 36-month prediction interval, while the diagnosis and medication models have lower AUROCs of 0.787 (95% CI: 0.776-0.791) and 0.746 (95% CI: 0.741-0.751) respectively (Figure 2A). As expected, the scores of models trained with data exclusion are lower when compared to the model without exclusion (see Supplementary), which can be attributed to the absence of quasi-symptoms.

**Figure 2.**
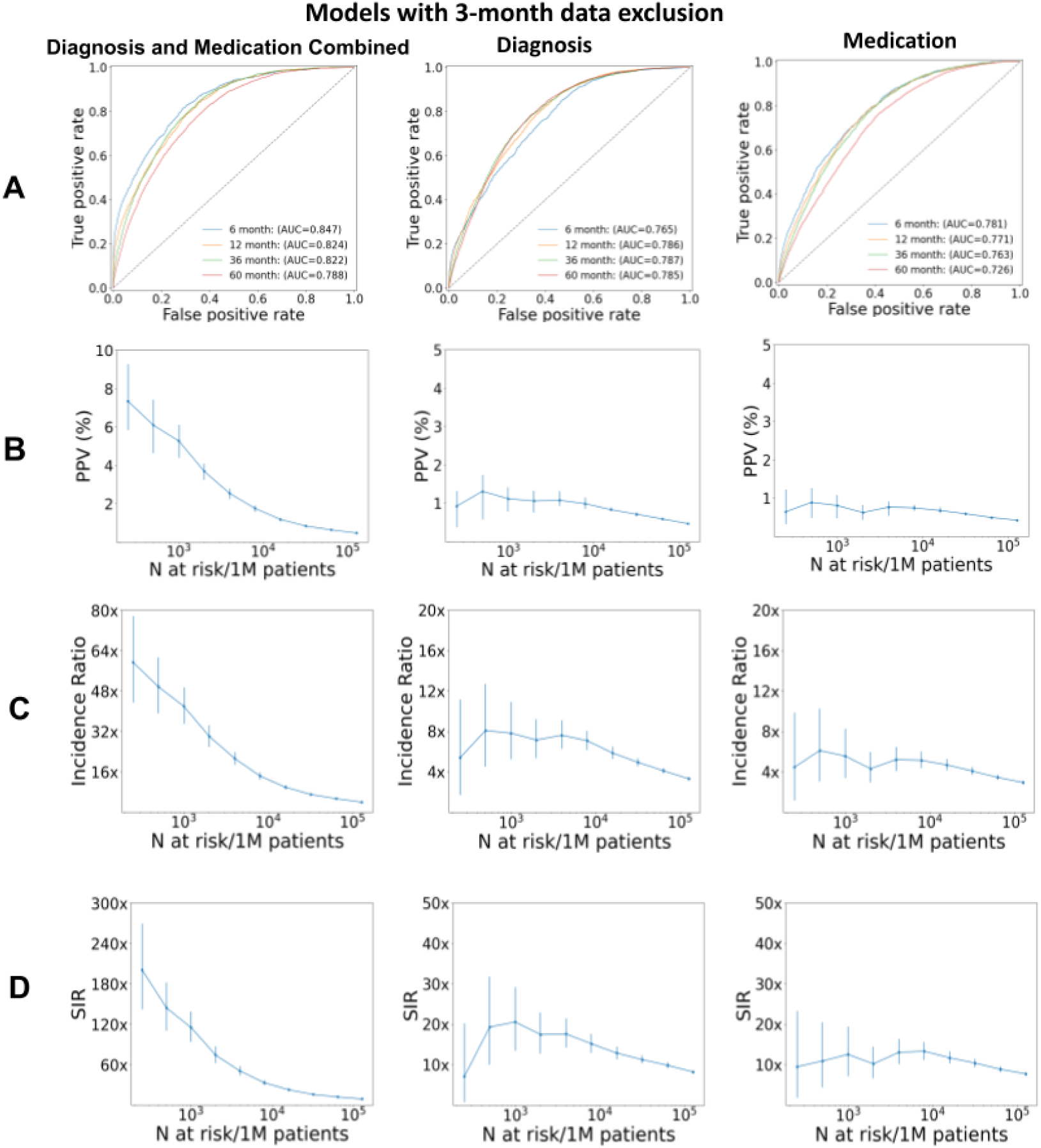
Comparison of the performance of model trained with a three-month data exclusion window for the 36-month prediction interval: diagnostic data only, medication data only, and combination of both. Left: model trained on the combination of diagnosis and medication data. Middle: trained on diagnosis data only. Right: model trained on medication data only. (A) ROC performance curve and area under the curve (AUC) for different prediction time intervals. (B) Positive Predictive Value (PPV) and (C) Incidence ratio and (D) Standardized incidence ratio (SIR) for the N highest-risk patients out of 1 million patients. The curve can be used to choose a decision threshold for realistic implementations.

With our focus on highest risk patients to be nominated for moderate-size affordable surveillance programs, we evaluate model performance (computed on the withheld test set) as a function of N highest risk patients out of 1 million real-world all-comers. The standard incidence ratio (SIR), which quantifies model performance relative to a simple model based only on incidence population statistics for a given gender and age bracket, decreases as one goes down the list of N patients ranked by predicted risk (Figure 2C). The curve reflects the tendency of generally higher accuracy at smaller N (except at very small N where small sample size results in higher uncertainty intervals). The curve can be used to make a particular choice of decision threshold such that N represents a conservative choice that minimizes false positives and optimizes performance by restricting the number N of high-risk patients nominated for surveillance. In a real-world application, the N-threshold should be selected where the confidence interval is low and stable, ensuring a balance between the cost and practical requirements of a clinical surveillance program targeting the highest-risk individuals.

To address the question of when in a prediction interval cancer is likely to occur, we evaluate, as an example, the actual time to cancer (in the test set) for N=1000 highest risk patients for the (Figure 3). In this example, 37% of the highest risk patients have cancer occurrence after more than 100 days. This kind of evaluation can be a guide to choice of tests and follow-up repeat visit schedules in the design of surveillance programs.

**Figure 3.**
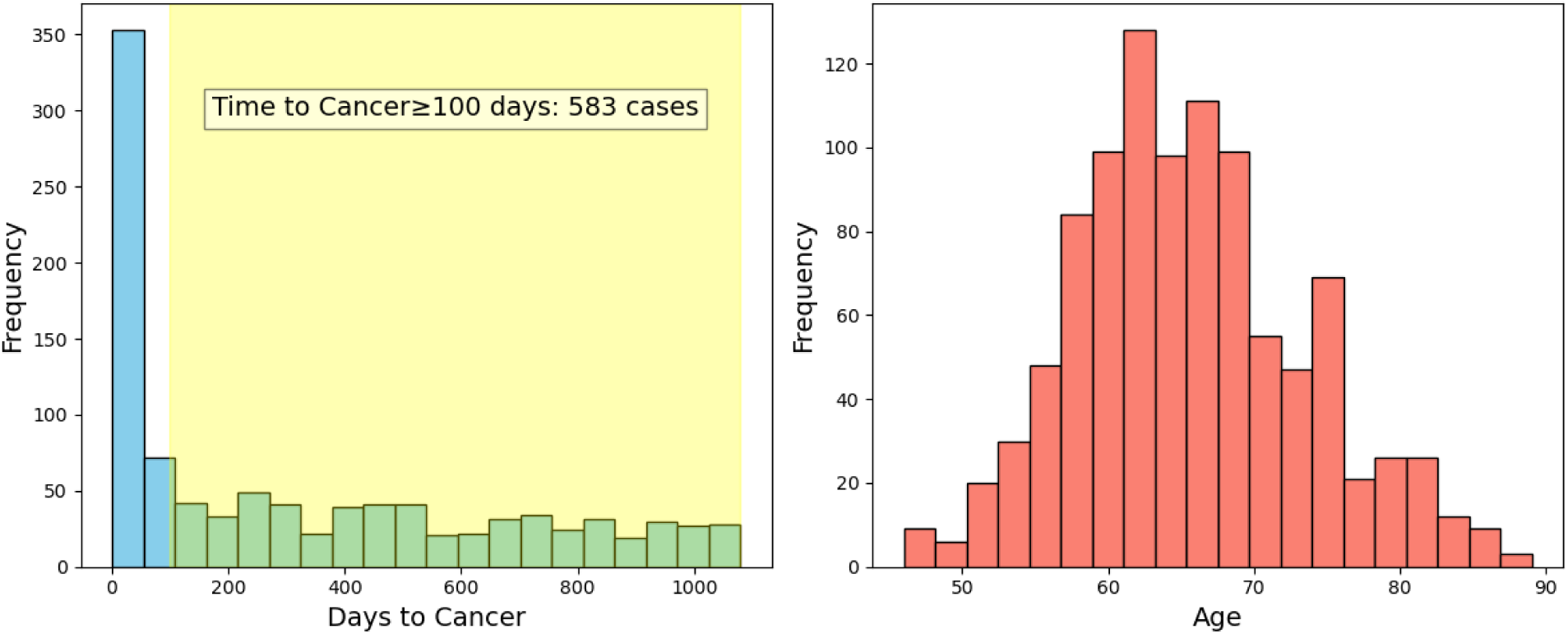
Characteristics of highest-risk subpopulation using a model trained with a three-month data exclusion window for the 36-month prediction interval (N=1000) (Left) Patients who were predicted to be at **high risk** and eventually **got cancer** are sampled to determine the time difference between the end of the disease trajectory (time point of prediction) and the actual cancer diagnosis (x-axis). y-axis: frequency of patients in the respective time gap. The frequency at larger time differences (yellow) indicates to what extent the model can correctly predict cancer diagnosis at larger time intervals before the actual diagnosis. **(Right)** Age distribution in the sampled high-risk cohort indicates good coverage.

### Interpretation of most predictive features

To highlight the most relevant features driving the prediction of the risk model (attribution analysis), we applied the Integrated Gradients (IG)(35) method to a set of high-risk patient trajectories. Features consisting of ICD codes and medications were ranked by their IG scores, with the top 10% identified as most predictive (details in supplementary; list in Table 2).

**Table 1.** Patient records in the US-VA dataset for model training and evaluation. US-VA data includes patient visits in both inpatient and outpatient settings across the nation. (A) The dataset for model development has 18K cancer cases and a randomly selected set of 193K controls. (B) The dataset for model evaluation (also called held-out dataset in Methods) has a subset of 1300 cancer cases and, to reflect the balance in real-world data, 986K controls. Q1: first quantile and Q3: third quantile.

| (A) | Overall | Non-cancer | Cancer |
| --- | --- | --- | --- |
| **n (number of patients)** | 210848 | 192722 | 18126 |
| **By gender, n (%)** |  |  |  |
| Female | 28071 (13.3) | 27566 (14.3) | 505 (2.8) |
| Male | 182774 (86.7) | 165153 (85.7) | 17621 (97.2) |
| **By race, n (%)** |  |  |  |
| Asia/Pacific Islander | 3066 (2.1) | 2917 (2.2) | 149 (1.0) |
| Black | 25708 (17.3) | 22670 (16.9) | 3038 (20.6) |
| Native | 1159 (0.8) | 1062 (0.8) | 97 (0.7) |
| White | 119061 (79.9) | 107582 (80.1) | 11479 (77.8) |
| **Trajectories-ICD diagnoses** |  |  |  |
| Lengths (# events), median [Q1,Q3] | 91.0 [15.0,298.0] | 82.0 [13.0,283.0] | 195.0 [73.0,433.0] |
| Time (years), median [Q1,Q3] | 7.0 [2.0,14.0] | 8.0 [2.0,14.0] | 7.0 [3.0,12.0] |
| **Trajectories-medications** |  |  |  |
| Number of prescriptions median (# events) [Q1,Q3] | 50.0 [14.0,129.0] | 48.0 [13.0,126.0] | 73.0 [29.0,152.0] |
| Time (years), median [Q1,Q3] | 8.0 [3.0,14.0] | 8.0 [3.0,15.0] | 7.0 [2.0,12.0] |
| **(B)** | **Overall** | **Non-cancer** | **Cancer** |
| **n (number of patients)** | 987693 | 986393 | 1300 |
| **By gender, n (%)** |  |  |  |
| Female | 141224 (14.3) | 141179 (14.3) | 45 (3.5) |
| Male | 846468 (85.7) | 845213 (85.7) | 1255 (96.5) |
| **By race, n (%)** |  |  |  |
| Asia/Pacific Islander | 15544 (2.3) | 15534 (2.3) | 10 (0.9) |
| Black | 114733 (16.6) | 114512 (16.6) | 221 (20.6) |
| Native | 5449 (0.8) | 5445 (0.8) | 4 (0.4) |
| White | 553383 (80.3) | 552544 (80.3) | 839 (78.1) |
| **Trajectories-ICD diagnoses** |  |  |  |
| Lengths (# events), median [Q1,Q3] | 83.0 [13.0,282.0] | 82.0 [13.0,282.0] | 207.0 [78.0,458.5] |
| Time (years), median [Q1,Q3] | 8.0 [2.0,14.0] | 8.0 [2.0,14.0] | 7.0 [3.0,12.0] |
| **Trajectories-medications** |  |  |  |
| Number of prescriptions median (# events) [Q1,Q3] | 48.0 [13.0,126.0] | 48.0 [13.0,126.0] | 78.0 [31.0,167.0] |
| Time (years), median [Q1,Q3] | 8.0 [3.0,15.0] | 8.0 [3.0,15.0] | 7.0 [2.0,12.0] |

**Table 2.** Top ten most predictive diagnosis and medication codes using a model trained with a three-month data exclusion window for the 36-month prediction interval. We conducted an attribution analysis on high-risk patients and list the most predictive diagnosis (left) and medication (right) codes for high-risk patients as identified by our model.

|  | Diagnosis Codes |  | Medication Codes |
| --- | --- | --- | --- |
| 1 | Essential (primary) hypertension | 1 | Lisinopril |
| 2 | Non-insulin-dependent diabetes mellitus, Insulin-dependent diabetes mellitus, Other specified diabetes mellitus | 2 | Simvastatin |
| 3 | Disorders of lipoprotein metabolism and other lipidaemias, Other metabolic disorders, Disorders of glycoprotein metabolism | 3 | Hydrochlorothiazide |
| 4 | Acute pancreatitis | 4 | Aspirin |
| 5 | Chronic ischaemic heart disease | 5 | Metformin |
| 6 | Psoriasis, Parapsoriasis, Pityriasis rosea Other papulosquamous disorders, Other dermatitis | 6 | Hydrocortisone |
| 7 | Benign neoplasm of other and ill-defined parts of digestive system | 7 | Nitroglycerin |
| 8 | Other chronic obstructive pulmonary disease | 8 | Docusate |
| 9 | Polyarthrosis, Other arthrosis, Coxarthrosis | 9 | Insulin |
| 10 | Glaucoma, Other congenital malformations of eye | 10 | Vitamin B12 |

Essential hypertension and diabetes have widely been associated with cancer risk (36), while lipoprotein metabolism disorders link to disrupted lipid metabolism (37). Conditions such as acute pancreatitis (38) and chronic inflammatory conditions such as ischemic heart disease as well as chronic obstructive pulmonary disease are linked to sustained inflammation (39). These diagnoses include several metabolic, cardiovascular, and inflammatory conditions which are known to be associated with high risk of pancreatic cancer.

Metformin, prescribed medication for type 2 diabetes, and insulin, used for blood glucose regulation, both are associated with elevated glucose levels known to increase risk of cancer (40,41). An association between the initiation of antidiabetic and anticoagulant medications and the diagnosis of pancreatic cancer in a two year pre-diagnosis time window was also reported in a study of controlled cohorts of nurses and health professionals (42). Aspirin and simvastatin, prescribed in cardiovascular or chronic inflammation conditions, may influence cancer risk through their anti-inflammatory effects (43,44). These medications likely indicate underlying metabolic or inflammatory conditions that may increase the likelihood of future cancer development, suggesting their role as markers of disease progression rather than direct causative agents.

These interpretations are to be considered with caution, as the IG method does not provide isolated effects of individual features; instead, it measures the influence of each feature change within the context of other features in the input vector. This implies that the predictive power of features emerges from their interdependencies rather than their independent contributions.

### Evaluation in subpopulations

AI models can be biased towards specific subpopulations based on the distribution of samples from different groups within the training dataset (45). For instance, if the training data predominantly includes samples from one demographic group, the model is likely to perform better when evaluated on that specific group. Investigating such biases is important for assessing whether a model performs comparably across all subgroups.

We, therefore, stratified the evaluation dataset and validated the model on subpopulations defined by race and sex. The AUROC for white patients in a 12-month prediction window is 0.901 (95%CI: 0.897-0.904), compared to 0.870 (95%CI: 0.860-0.880) for black patients. For male patients it is 0.892 (95%CI: 0.888-0.896), compared to 0.900 (95%CI: 0.883-0.913) for female patients. Moreover, when examining the 1,000 high-risk cohorts, we observed that the SIR for white patients is higher than for black patients, and the SIR for male patients is higher than for female patients. There are more white patients and more male patients in the database (Table 1), contributing to the observed performance disparities for the models trained on all patients. These gaps highlight the model’s bias plausibly resulting from the uneven representation of subpopulations in the dataset (Figure 4).

**Figure 4.**
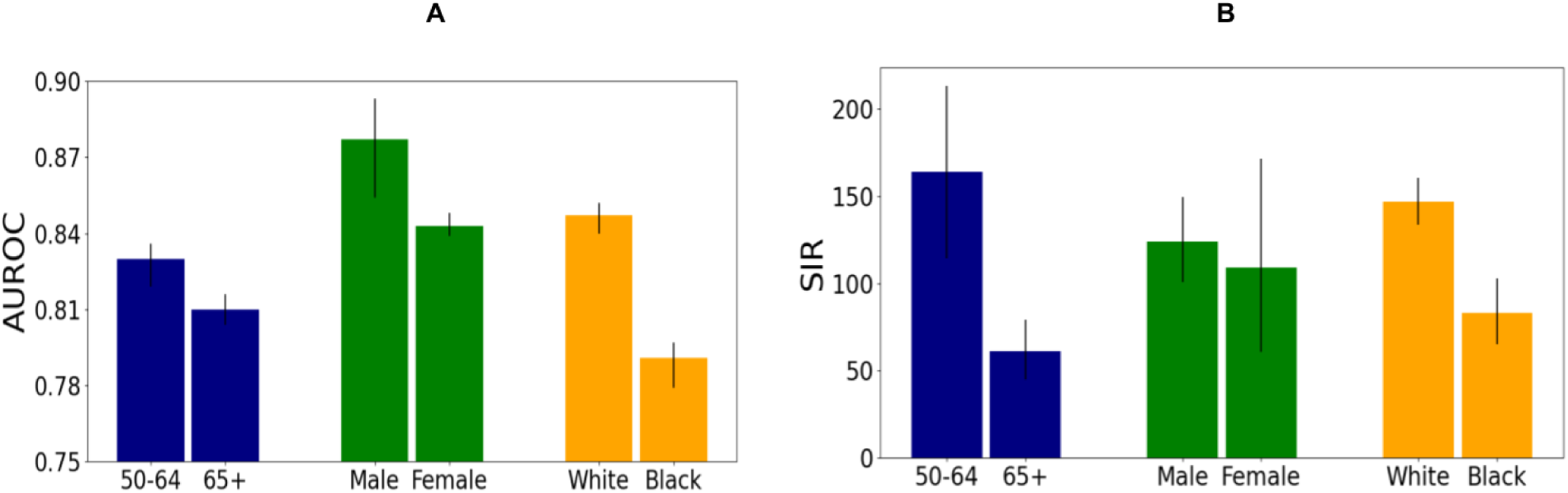
Evaluation of prediction performance in subpopulations using a model trained with a three-month data exclusion window for the 36-month prediction interval. **(A)** AUROC and **(B)** SIR, stratified by race, gender, and age. Notably, the SIR score is higher for the white subpopulation, as well as for male patients, and individuals in the 50-64 age group. The observed differences in performance across subpopulation groups can be attributed to biases in the dataset, as it consists of a higher proportion of white patients compared to black patients, more male patients compared to female patients, and a greater number of individuals in the 50-64 age group compared to 65+ age group, as detailed in Table 1.

**Figure 5.**
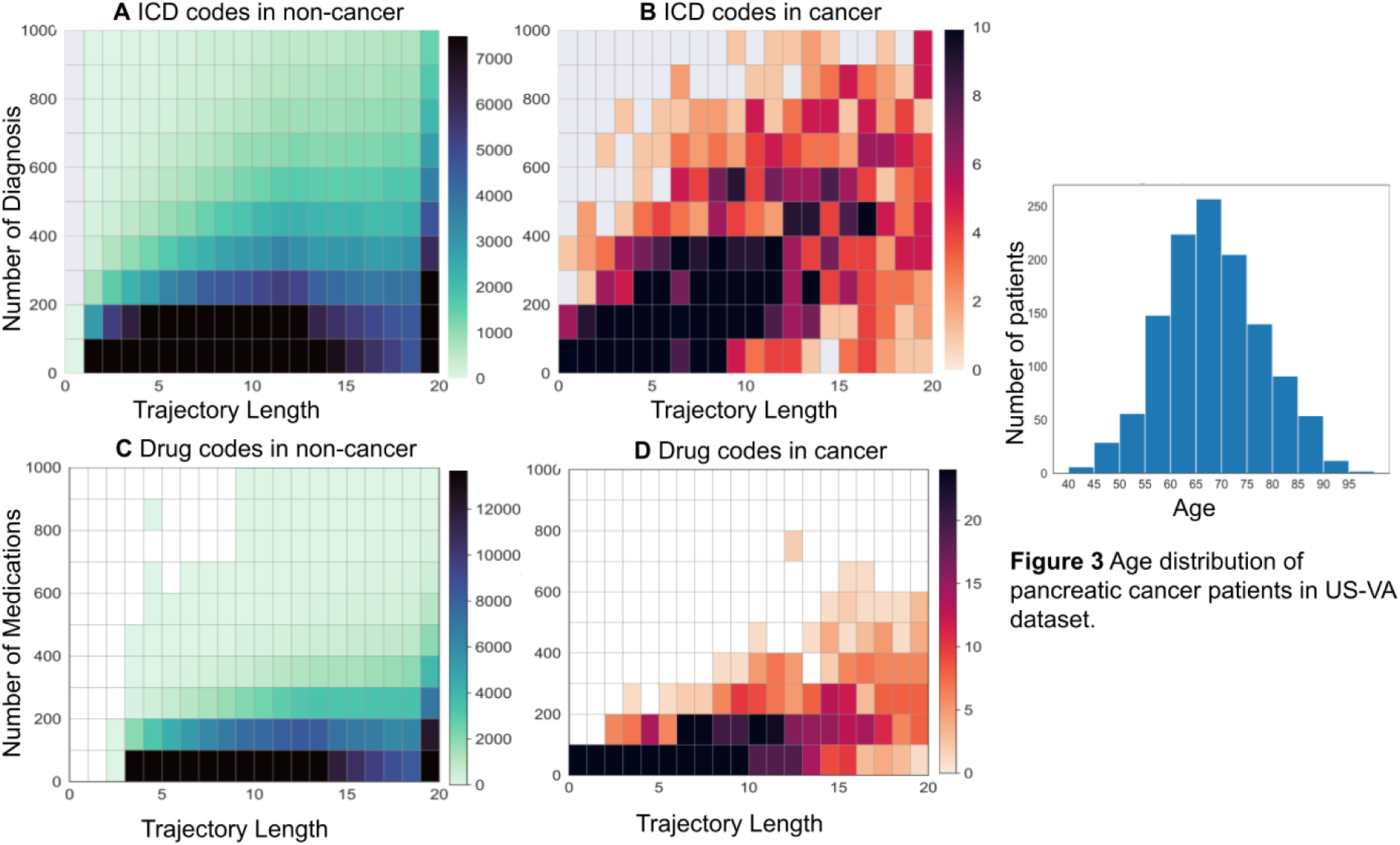
The density of medications and diagnosis codes for different data trajectory lengths. Cancer patients (A,C), and non-cancer patients (B, D). The intensity of color is a number of patients in a bin. For non-cancer patients, patients with less than two years of follow-up were removed.

## Methods

### Datasets for model development and evaluation

We use the US-VA database to construct training and evaluation datasets, based on a total of 15,926,415 patients, including 19,426 cases of pancreatic cancer. Overall data statistics including data splits used for model development and evaluation are in Table 1. We use each patient’s time-based trajectory of International Classification of Diseases (ICD) codes for diagnoses and active ingredients from prescription medications. Consistent with the approach in earlier work (34), we restrict ICD codes to three-character categories within the ICD hierarchy to focus on fairly broad diagnostic groups. Timestamps associated with the diagnosis or medication event codes are incorporated in the model as positional encodings to account for the temporal aspect of patients’ clinical histories.

To train the AI model, we subsample trajectories of fixed lengths for each patient, starting at the first recorded event and ending at various endpoints. This subsampling approach ensures a more comprehensive modeling of patient risk, as it captures periods when specific symptoms develop. As a result, the same patient can have trajectories representing variations in cancer risk over time. These variations can occur for a number of reasons such as changes in lifestyle, disease states or the use of specific medications. This approach allows us to interpret each trajectory as a realistic and dynamic representation of a patient’s health over time.

### Evaluation of prediction performance

Using different data types, including patient ICD codes, medication usage and their combination, we trained three models and then evaluated their performance using four metrics: AUROC, positive predictive value (PPV), incidence ratio (IR) and standardized incidence ratio (SIR). Further details on each metric are in the Supplementary. The SIR is particularly relevant for developing a realistic surveillance program, as it compares the model’s predictions to standardized incidence rates from population-wide studies conditioned on age and gender. A SIR of 1 indicates a model prediction is equivalent to frequency estimates based on population-wide studies, while higher values indicate better predictive performance. Considering the high cost of surveillance programs, these are currently most realistic for a relatively small number of patients at a very high risk of pancreatic cancer. We, therefore, select the top 1,000 high-risk patients from a cohort of 1 million and report the SIR scores for different models.

### Risk prediction model

The architecture of the risk prediction model is based on the framework described earlier (34). The earlier implementation only uses diagnostic codes. The current model takes as input combined trajectories of medication and diagnostic codes, along with their respective timestamps, ending at the time of prediction. Initially, all events in a given trajectory of clinical data items are encoded into a fixed-length representation using an embedding layer. The timestamp information is incorporated in form of relative positional encoding of events, which, when combined with the embedding, provides a time-dependent encoding of trajectories. These encodings are then input into a Transformer model which uses a self-attention mechanism to learn interdependencies between all codes within a trajectory, both diagnostic and medication. The Transformer’s output is aggregated along the time dimension to produce a fixed-length latent representation of a given input trajectory. This representation is subsequently fed into a supervised risk regression model, which predicts the probability of cancer occurrence within five time intervals: 0-3, 0-6, 0-12, 0-36, and 0-60 months. Further details are in the Supplementary.

## Discussion

### Combination model trained on diagnostic and medication trajectories

This work highlights the information value of combining diagnosis and medication data in patients’ health histories using a Transformer neural network for risk prediction of pancreatic cancer. Medication data carries information complementary to diagnosis codes, and helps address the hard-to-detect and hard-to-correct upcoding of diagnosis codes, as medication codes are less likely to be influenced by finance-motivated billing practices (23). In future, application of natural language processing of clinical notes to filter diagnosis codes and ML-learned feature selection may be useful to mitigate billing related noise in EHR datasets. In addition, additional patient data such as blood test results and, in future, widely available genetic information will probably further enrich the model’s ability to learn from the most informative combinations of risk factors in a real-world population.

### Sequential modeling of EHR trajectories

By using a sequential model, the AI tool makes use of temporal dependencies between health events. Our results indicate that a sequential model, as opposed to a ‘bag-of-events’ model, can capture temporal contributions of and informational synergy between disease and medication events, leading to improved performance in predicting cancer risk compared to non-sequential models. (Supplementary Figure 2S).

### Focus on large real-world patient data and on the highest risk bracket

There is an informational trade-off between large real-world EHR datasets and specialized data from small specifically recruited cohorts. This study focuses on large real-world patient datasets in contrast to pioneering earlier prediction models in smaller controlled cohort studies, which have developed risk prediction models using carefully acquired well-defined risk factors such as family history, genetic markers, unintended weight loss, and systematic changes in fasting glucose levels (6–11). However, such valuable information is not generally available in the EHRs of all-comers in large populations. To cast a wider net, this study used EHRs from real-world populations based only on generally available information in health records, such as diagnoses and prescriptions.

In a trade-off that balances cost and benefit of surveillance programs, we focus on the selection of high-risk cohorts, which is only a small fraction of the large total population: patients at very high risk for the cancer with the highest probability of accurate prediction (lowest false positive prediction rate), for whom expensive screening tests are justified and affordable. For example, from all patients in a hospital system aged 50 years or older, out of 1 million patients on whose EHRs the prediction tool is applied, we propose to identify a subpopulation of a few thousand highest-risk patients and focus on a 12-month prediction interval. E.g., for 1000 patients at highest risk, the result of SIR=115 for a model trained with a 3-month exclusion window (not using data from a 3 month interval before cancer occurrence) quantifies how much better the AI tool is in identifying high-risk patients than usage of a background model that uses population wide incidence rates, only using age and gender information. The tool thus may have concrete utility as a decision support tool aiding clinicians in an early detection program in a large hospital system.

### Prediction performance in subpopulations

We have also evaluated the prediction performance of the inclusively trained model in different race groups and highlight potential biases to be addressed in future work. Our retrospective (on a withheld test set) evaluation of the PDAC risk prediction model across different race groups highlights the potential impact of inherent biases in data distribution. Specifically, the model trained on the entire population had a lower AUROC score of 0.82 for the black ethnic group compared to 0.85 for the White ethnic group; a lower score of 0.86 for male subpopulation compared to 0.88 for female subpopulation; and higher score of 0.86 for patients aged 50-64 compared to 0.82 for age 65+ patients (Fig. 4). These disparities in performance can plausibly be attributed to inherent differences in the training data distribution, which leads to biases in prediction performance. For instance, there are more white than black patients in the EHR database (Table 2) so that disease and medication data items corresponding to white are more likely to be reflected in the parameters of the trained model. In addition, differences in the type of EHR data items in each subpopulation often reflect existing disparities within society, especially socioeconomic status (SES) (46). This is because social determinants of health (SDOH), such as access to health insurance and healthcare services, are typically influenced by financial income (47). Factors such as historical demographics, data collection procedures, and access to healthcare services can directly influence the composition of population-specific data in a healthcare system and lead to underrepresentation or overrepresentation of specific populations in the dataset.

### Addressing disparities in performance across demographics groups

While differences in the performance of cancer risk prediction methods in subpopulations have been noted earlier (3), addressing these as part of an improved ML process is an open problem, which we have not addressed in this study. A relevant future task would be to evaluate models of this type on groups with diverse socio-economic status (SES), perhaps by the use of residential information, such as postal zip codes, if this information is available. With sufficient population structure data, one can train a separate model for each ethnic or socioeconomic group in a particular health system. Access to such data could be obtained within a unified national healthcare system, which is not currently available in the US, or by federated learning procedures with software access to data behind institutional firewalls, which are technically feasible but difficult to negotiate. The key aim is to remove bias in the trained model(s) and achieve equal accuracy in spite of unequal risk. We anticipate that future work will achieve this aim in support of more equitable access to early detection and prevention programs.

### Stratified surveillance programs

The proposed PDAC risk model, which predicts cancer risk probabilities over various time intervals after risk assessment, offers a promising approach to differentially screen individuals based on their risk profiles. Such stratification can be either binary (at risk or not), or more granular using the real-number probabilities to stratify patients into, e.g., very high, high, medium, and low-risk groups. One can also determine thresholds in the probability values as decision parameters for various screening actions. Differential action can imply different time intervals for repeat screening visits, different procedures such as CT scans, MRI scans, endoscopic ultrasound or different advanced blood tests, e.g., for sequence variants, methylation or fragments in cell free DNA or for serum proteins, once these are generally accepted. Cost of the program and value of the kind of information desired would be decision parameters. Suggesting details of stratified screening programs is beyond the scope of this study. These are likely to be worked out by clinicians in consultation with developers of detection technologies, machine learning experts and hospital administrators. Careful definition and prospective evaluation of stratified efficient surveillance programs is likely to lower the barrier for general acceptance in clinical practice.

## Outlook

The proposed AI-based risk prediction tool can be an important component of a clinical cancer management program, identifying only high-risk individuals drawn from a large, real-world population for an initial moderate-size lower-cost targeted surveillance program. By focusing on such high-risk cohorts, one can achieve a reduction in predicted false positives and a reasonably precise identification of pre-cancerous and early-stage cases. Lowering screening costs helps ease the healthcare system’s burden, while early identification facilitates preventive measures, such as lifestyle modifications, that may delay or prevent pancreatic cancer onset. An AI-driven surveillance program—combining prediction, detection, and treatment within a clinical collaboration—provides a promising framework for equitable, effective cancer surveillance across diverse healthcare environments.

## Contributors

Author contributions: concepts C.Z., D.R., A.K., D.S.M, N.R.F, and C.S.; computation C.Z., D.R., and A.K.; supervision D.S.M, N.R.F, and C.S.; manuscript C.Z., D.R., A.K., D.S.M, N.R.F, and C.S. By mutual agreement, joint first authors can list the article as Name et al.

## Data Availability

Due to privacy regulations, the data used in this study is protected. Those who wish to access these data should contact the VA and obtain the necessary Institutional Review Board (IRB) approvals, in accordance with VA policies.

## Acknowledgements

NONE

